# Mapping Climate Change’s Impact on Cholera Infection Risk in Bangladesh

**DOI:** 10.1101/2022.06.09.22276227

**Authors:** Sophia E. Kruger, Paul A. Lorah, Kenichi W. Okamoto

## Abstract

Several studies have investigated how *Vibrio cholerae* infection risk changes with increased rainfall, temperature, and water pH levels for coastal Bangladesh, which experiences seasonal surges in cholera infections associated with heavy rainfall events. While coastal environmental conditions are understood to influence *V. cholerae* propagation within brackish waters and transmission to and within human populations, it remains unknown how changing climate regimes impact the risk for cholera infection throughout Bangladesh. To address this, we developed a random forest species distribution model to predict the occurrence probability of cholera incidence within Bangladesh for 2015 and 2050. Using R, our random forest model was trained on cholera incidence data and spatial environmental raster data at a resolution of 250 square meters. This model was then predicted to environmental data for the training data year (2015) and for 2050. We interfaced R with ArcGIS to develop risk maps for cholera infection for the years 2015 and 2050, proxying infection risk with cholera occurrence probability predicted by the model. The best-fitting model predicted cholera occurrence given elevation and distance to water. We find that although cells of high risk cluster along the coastline predominantly in 2015, by 2050 high-risk areas expand from the coast to inland Bangladesh with all but the northwestern district of Rangpur seeing increased clusters around surface water. Mapping the geographic distribution of cholera infections given projected environmental conditions provides a valuable tool for guiding proactive public health policy tailored to areas most at risk of future disease outbreaks.

## Introduction

Cholera, a waterborne bacterial disease that causes severe diarrhea and dehydration in humans, remains a significant threat to global health. Despite proposed efforts to reduce global cholera mortality by 90% by 2030 (World Health Organization, 2017), researchers estimate that between 1.3 million and 4 million cholera cases occur annually, with an estimated 21,000 to 143,000 deaths (Ali et al., 2015).

The etiological agent *Vibrio cholerae* resides in coastal brackish water and riverine habitats and is typically seeded along coastlines (Escobar et al., 2015). Among many proposed hosts, vectors, and reservoirs of infection, zooplankton remain the largest known environmental reservoir of *V. cholerae* (Vezzulli et al., 2010). Consumption of seafood or water contaminated with an infective dose of free-floating *V. cholerae* or *V. cholerae-*harboring zooplankton causes human infections, while infection may also occur through fecal-oral transmission between human hosts. Such transmission pathways are influenced by environmental conditions in waterbodies that favor bacterial growth (Lemaitre et al., 2019). Changes to such waterbodies influence the epidemiology and ecology of *V. cholerae* by altering bacterial reproduction, transmission, and exposure risks.

Climatic conditions, such as rainfall and sea surface temperature, thus drive epidemiological risk, with warmer, wetter environments increasing the likelihood of disease transmission and infection (Christaki et al., 2020). Salinity, pH, and sea surface temperature have been shown to encourage bacterial growth and hence *V. cholerae* infection risk and endemicity in the Bay of Bengal (Islam et al., 2020).

However, future climate conditions can also promote increased infection risk in inland populations. Heavy rainfall events (e.g., El Niño and Southern Oscillation and summer monsoons) increase cholera infection risk by damaging sanitation systems and contaminating water sources with sewer spillage (Lemaitre et al., 2019; Moore et al., 2017; Koelle et al., 2005). Surface water contaminated with brackish coastal waters may also serve as sources of infection after flooding events (Khan et al., 2017). Cholera infection risk may also increase in periods of drought, during which reliance on scarce water sources increases the likelihood of contamination with *V. cholerae,* especially if human hygiene practices partake in waters used for drinking water (Hashizume et al., 2008).

Curbing widespread infection, mortality, and social disruption requires characterizing the epidemiological risk for cholera, which in turn depends on how regional weather, land-use practices, and climate conditions influence cholera epidemiology and ecology. Risk mapping, a method of associating risk values to explicit geographic areas, has become an effective tool for not only visualizing the spatial distribution of disease burden (i.e., risk) but also for guiding public health policy to reduce that burden (Peterson, 2014; Leta et al., 2018).

One approach to estimating risk across a landscape is to use non-mechanistic correlative models that predict infection risk given disease incidence data (e.g., disease presence/absence) and environmental covariates. Predicting risk under future environmental and climate scenarios is essential for disease surveillance and is a powerful tool in guiding proactive public health policy for areas most at risk of future disease outbreaks. Such a strategy is particularly critical in endemic areas, as pandemic strains of *V. cholerae* almost invariably emerge from endemic areas that seed epidemics abroad (Azman et al., 2020; Christaki et al., 2020).

Several studies have sought to predict risk for cholera infection given climate and weather differences via risk-mapping (Xu et al., 2013; Baker-Austin et al., 2013; Escobar et al., 2015; Khan et al., 2017; Lessler et al., 2018; and Azman et al., 2020). Most risk-mapping studies restrict their analyses to present climatic conditions or limit climate projections to coastal settings only. To our knowledge, no study to date integrates long-term climate projections into risk mapping, especially for inland populations of endemic countries notoriously affected by climate change. Such an analysis is critical to sustaining effective regional public health strategies over the medium and long term.

Here we construct risk maps for cholera infection for Bangladesh under current and future climate scenarios. We identify spatial environmental variables associated with human cholera infection and cholera incidence data from a detailed country-wide serosurvey study (Azman et al., 2020), and employ a fitted random forest model to predict the risk of infection across Bangladesh at a fine spatial resolution. Below, we characterize our analyses in greater detail.

## Materials and Methods

### (a) Cholera occurrence data

We used a serosurvey dataset described in Azman et al. (2020; anonymized data are publicly available at https://github.com/HopkinsIDD/Bangladesh-Cholera-Serosurvey) that identifies cholera prevalence within Bangladesh for 2015 for our disease presence data. Of the 2930 surveyed individuals, the 639 predicted positive cases constituted our model’s presence data while the predicted 2291 negative cases constituted absence data. The approximate coordinate location of each surveyed individual was also used by our model to extract values from our spatial covariates. Notably, multiple presence or background points may exist at the same coordinate location as serum samples were often taken from multiple individuals within the same household.

### (b) Spatial environmental data

To develop our model, we considered 13 spatial variables known to correlate with *V. cholerae* occurrence and case incidence and for which data were available for 2015 and 2050 (Table 1). Given our interest in predicting risk for the entirety of Bangladesh, we restricted our variables to those with values available for each cell in the extent used. Moreover, as *V. cholerae* can be found in semiaquatic and seasonally aquatic settings (Colwell, 1996; Islam et al., 2020), we excluded environmental variables describing aquatic environments only. All raster datasets were projected to the World Geodetic System 84 (WGS 84) projection, resampled to a 0.00214° (approximately 250-m^2^) resolution, and cropped to the extent of our study area using the ‘raster’ package version 3.4-13 in R (R Core Team, 2021; Hijmans and van Etten 2012; also, see Supplementary Material).

**Table 1.**
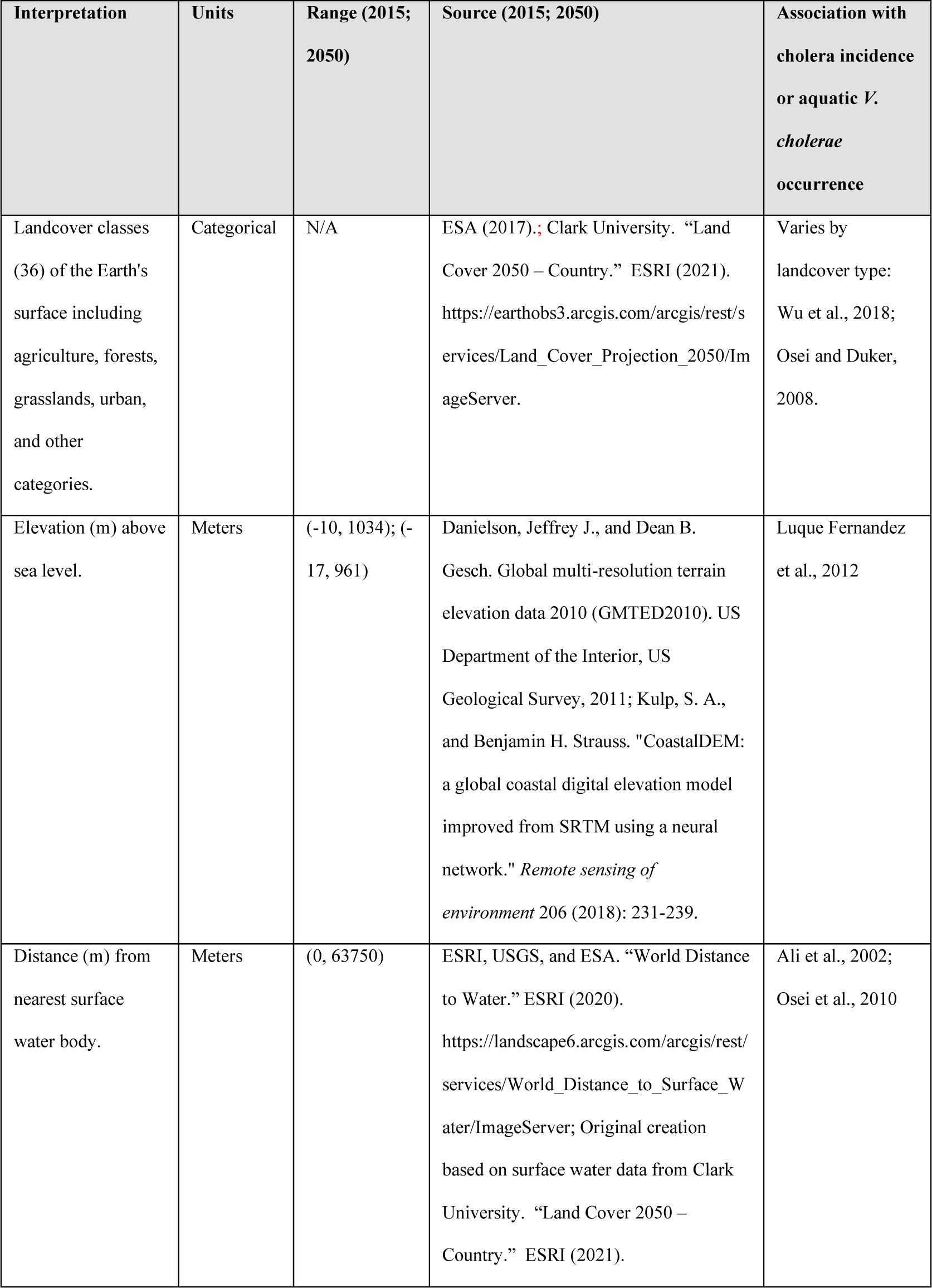
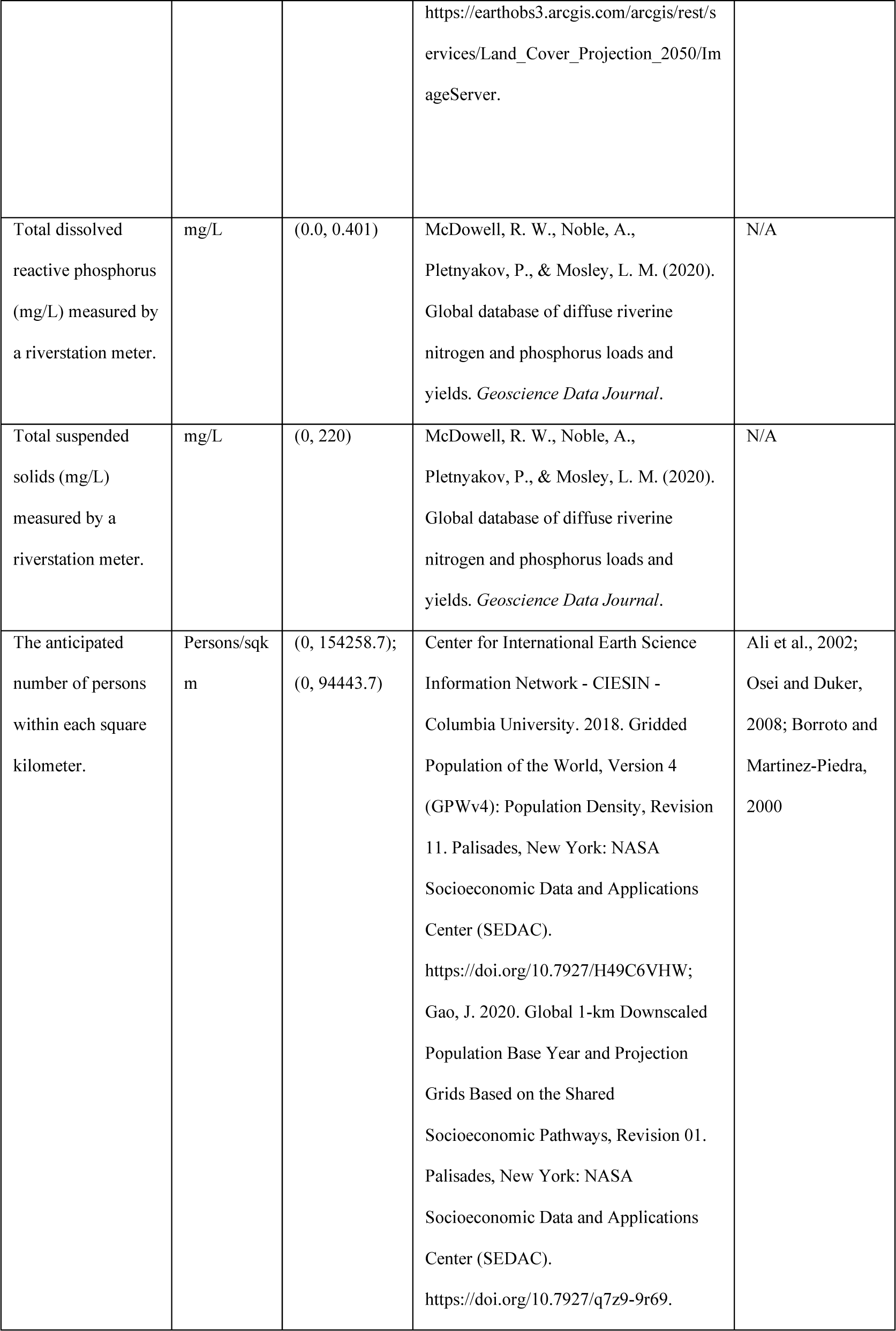
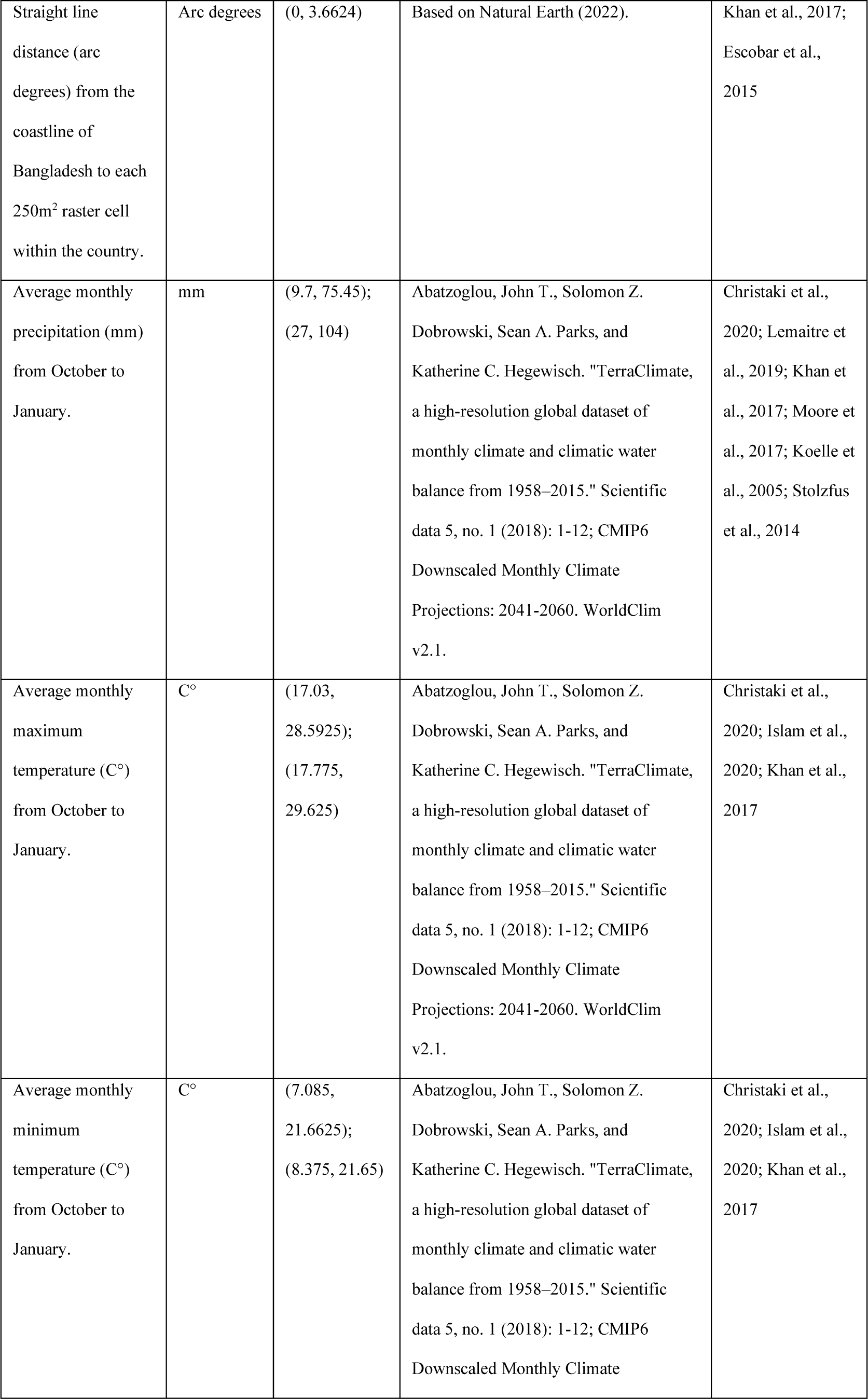
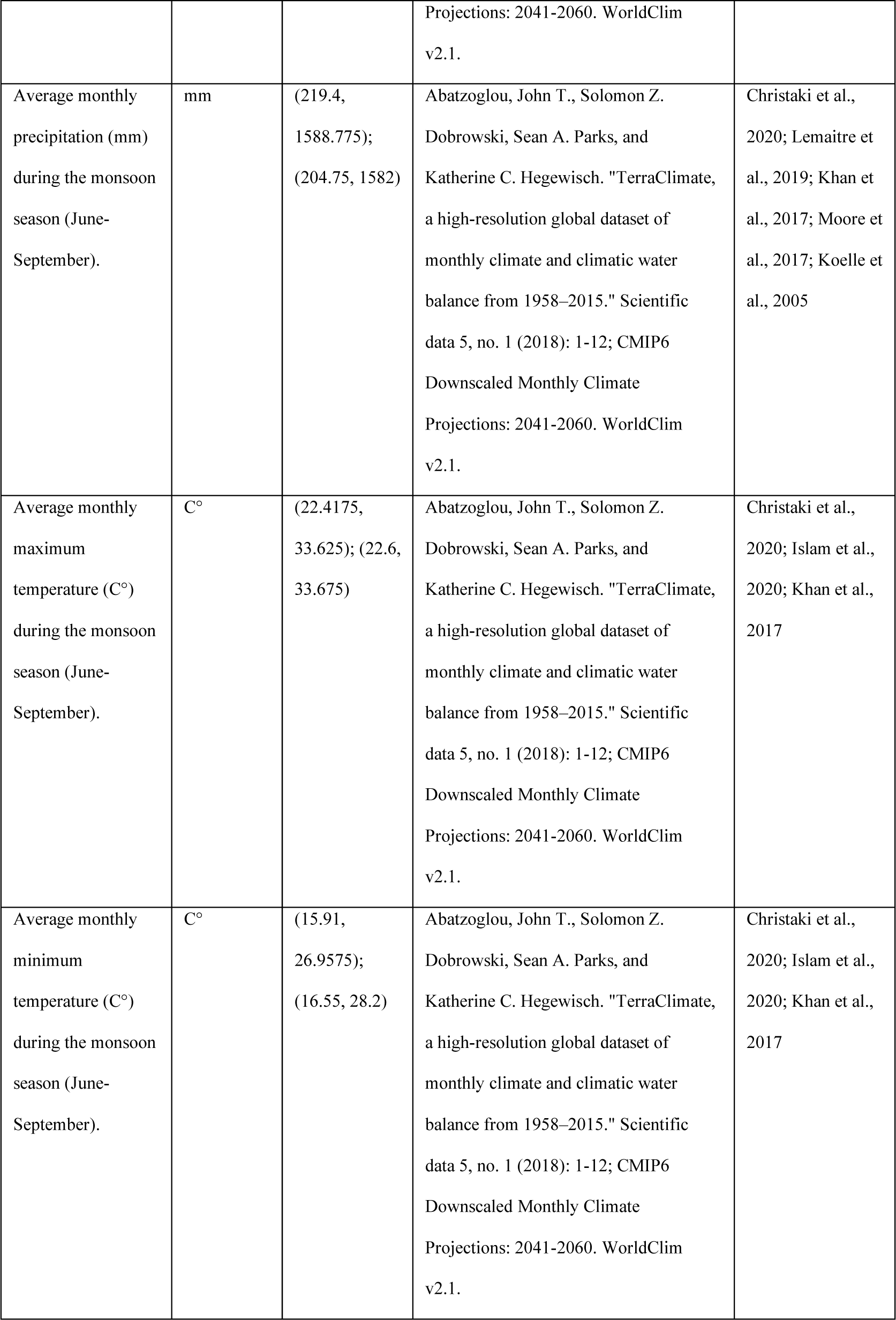
Description of the environmental variables considered in model building. Included, where available, are descriptions of each variable’s data (for 2015 and 2050), data sources, and references to known associations with cholera incidence.

### (c) Statistical analyses

We constructed a predictive model estimating cholera incidence in each 250-m^2^ raster cell in 2015 as a function of the spatial correlates using the presence-absence algorithm of the ‘randomForest’ package version 4.6-14 in R (Liaw and Wiener, 2002). Briefly, the random forest (RF) algorithm uses bootstrap aggregation and resampling to create an ensemble of lowly correlated decision trees that together classify each datapoint (Brieman, 2001; Muschelli and Varadhan, 2014).

To select the best-fitting model, we performed a stepwise model selection procedure using the variable importance measures from the RF model calibrated and evaluated with all covariates included. From this model, we selected the highest contributing variable to first create a univariate RF model using 80% of each sample group (i.e., presence and absence) as training data for model calibration and the remaining 20% for model evaluation. We ran the univariate models for 1000 iterations, computing the area under the curve (AUC) statistic from the receiver operating curve (ROC) generated for each run to create a 95% confidence interval of the AUC. From here, covariates were added individually to this model if the AUC confidence interval generated for the new model over 1000 iterations indicated improved predictive ability over the univariate model. For each iteration of the RF model, we used the algorithm’s default settings in R to perform a supervised classification.

Once the relevant variables were identified, we ran the best-fitting RF model from 2015 1000 times, training and evaluating the model of each iteration with the same 80% sample or 20% sample of presence-absence data, respectively. With each iteration, the model fitted to the 2015 data predicted cholera occurrence probabilities for 2050 for each 250-m^2^ cell. From these predictions, we constructed a mean, 2.5%-, and 97.5%-quantile rasterized map for each year by determining the mean, 2.5%-quantile, and 97-5%-quantile values for each cell within Bangladesh. Using the ‘arcgisbinding’ package in R, we interfaced ArcGIS Pro version 2.6.3 with R to transfer the raster maps generated in R to ArcGIS to ensure our rasters were of the appropriate resolution and extent (ESRI, 2019; ESRI, 2020). All code used in the analysis is publicly available on github (github.com/sophiakruger/cholera_risk) and released under the GNU Public License v.3 (Stallman, 2007).

## Results

### (a) Drivers of cholera infection risk

Our random forest classification model including all predictors (“the full model”) showed elevation as the most prominent predictor (*see* supplementary materials, Table S1). Thus, we began our stepwise model selection by sequentially adding other predictors to a univariate model with elevation as the sole predictor. The random forest classification model including elevation and distance to water as predictors increased the model’s predictive power compared to the full model and outperformed all other predictors that were added to the univariate model (Table 2). Model performance invariably declined when additional variables were added one at a time to the bivariate model (results not shown, *see* supplementary materials, Table S2). Generally, we find cholera infection risk increased with lower elevation and a shorter distance to the nearest surface water body (supplementary materials, figures S1 and S2).

**Table 2.** Summary of the top performing models obtained from our stepwise model comparison process. The area-under-the-curve (AUC) statistic identifies the bivariate model as the best-fitting model. The AUC confidence interval for the full model, univariate elevation model, and univariate distance-to-water model reflects the change of interval extremes (2.5% and 97.5% quantiles) from a null model (where AUC=0.50). The AUC confidence interval for the bivariate model reflects the increase in AUC from the mean AUC of the univariate elevation model.

|  | Model | AUC Conf. Int. |
| --- | --- | --- |
| **ΔAUC From Null (AUC=0.50)** | Full Model | (0.0852, 0.1045) |
|  | Elevation Only | (-0.0030, 0.0193) |
|  | Distance to Water Only | (0.0289, 0.0560) |
| **ΔAUC from Univariate Model** | Elevation + Distance to Water | (0.0841, 0.1088) |

**Figure 1.**
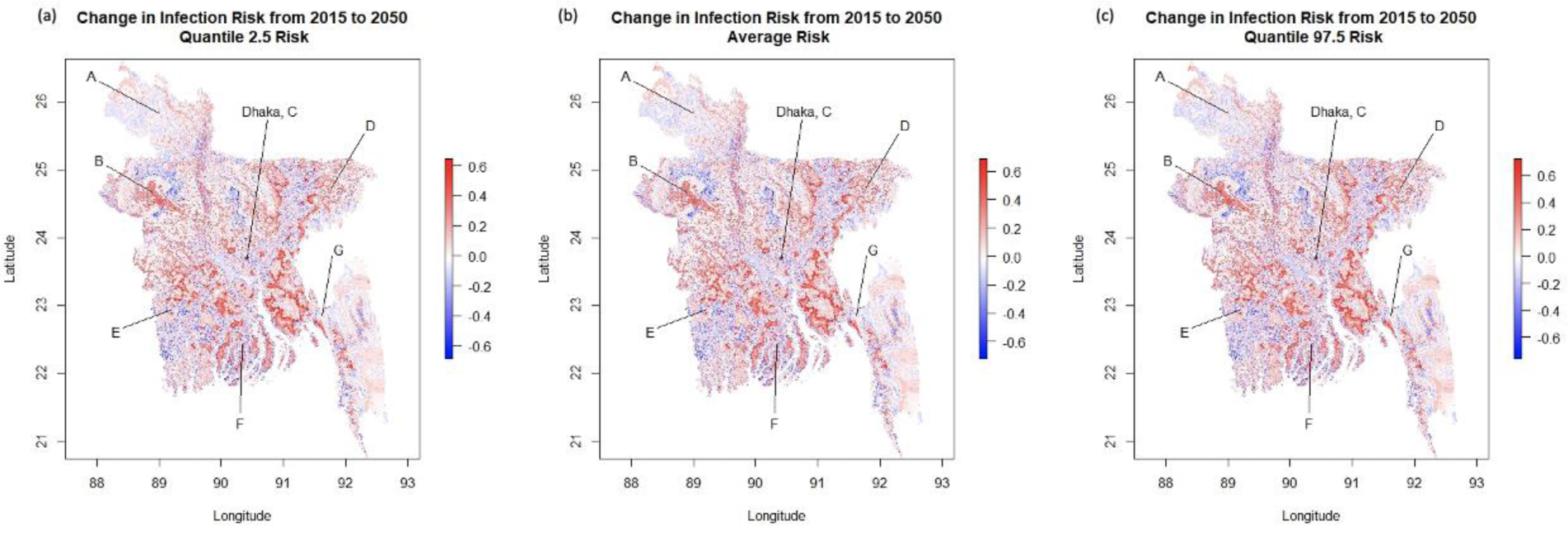
Change in occurrence probability (a proxy for infection risk) from 2015 to 2050 according to the bivariate random forest model with elevation and distance to water as predictors. The (a) 2.5% quantile, (b) mean, and (c) 97.5% quantile predicted values are shown with the districts of Rangpur (A), Rajshahi (B), Dhaka (C) with the capitol of Dhaka starred, Sylhet (D), Khulna (E), Barisal (F), and Chittagong (G). Supplementary figures S3 and S4 contain the underlying occurrence probabilities for each year.

**Figure 2.**
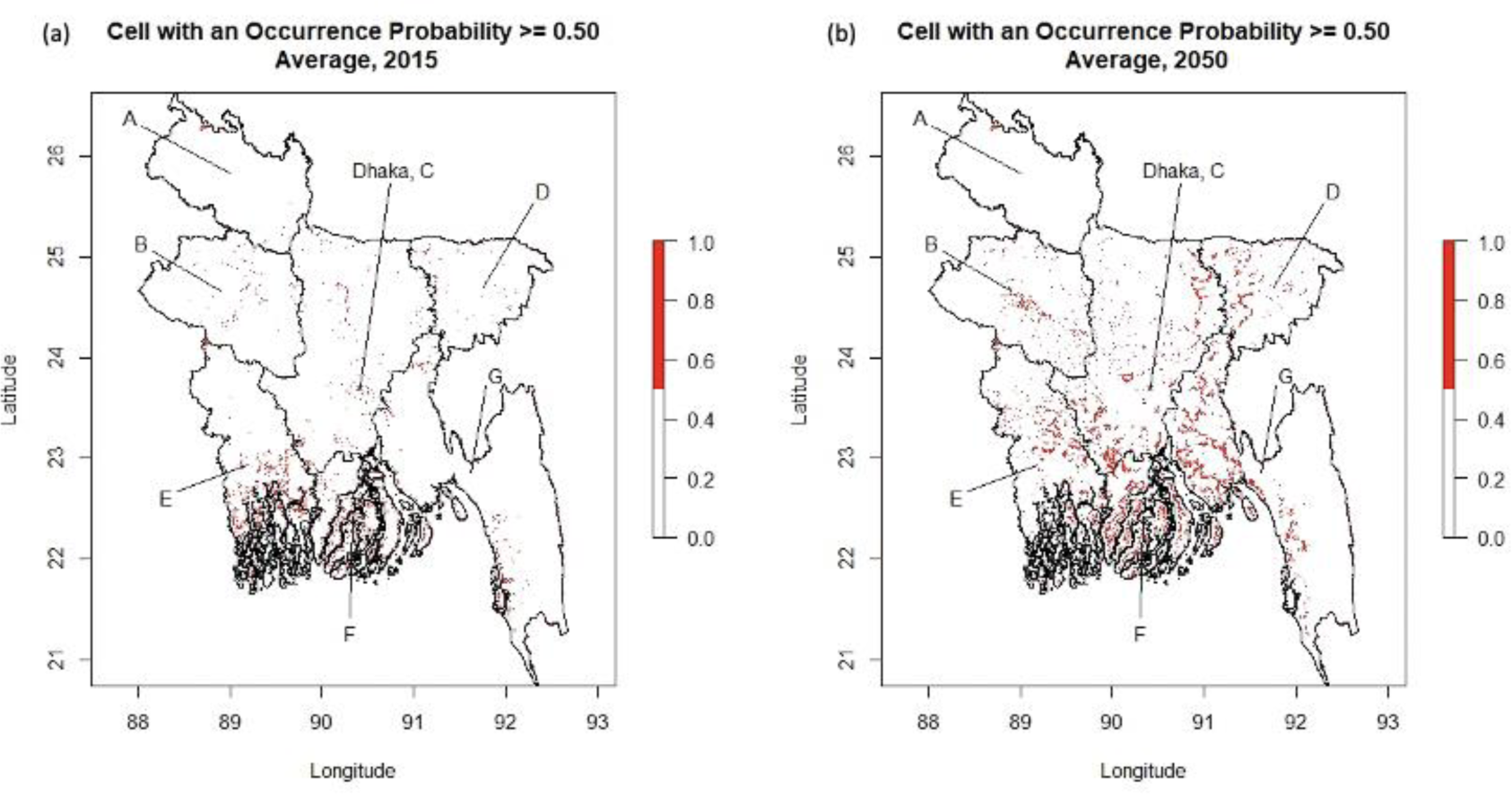
Average occurrence probability of 0.50 or greater for (a) 2015 and (b) 2050 according to the bivariate random forest model with elevation and distance to water as predictors. The districts of Rangpur (A), Rajshahi (B), Dhaka (C) with the capitol of Dhaka starred, Sylhet (D), Khulna (E), Barisal (F), and Chittagong (G) are highlighted. Supplementary figures S3 and S4 contain the underlying occurrence probabilities for the entirety of Bangladesh for each year.

### (b) Spatial predictions of cholera infection risk

We find that the distribution of cholera infection risk changes over time, with coastal and inland Bangladesh projected to experience an increase in cholera infection occurrence probability from 2015 to 2050 (Fig. 1(a)-1(c)). Even under the most conservative estimate for 2050, we find risk increases along tributaries, running inland from the coast (Fig. 1(a)). In 2015, cells with an average occurrence probability of 0.50 or greater cluster tightly along the coast of the Khulna and Barisal districts and are more widely distributed inland, though many follow the Padma River north into the district of Dhaka (Fig. 2(a)). Yet by 2050, clusters of cells with an occurrence probability of 0.50 or greater are predicted to increase inland in the districts of Khulna, Barisal, Chittagong, Rajshahi, Dhaka, and Sylhet (Fig. 2(b)). Notably, while an occurrence probability of 0.50 and greater cluster around major river systems along district boundaries in 2015, by 2050 these risk clusters expand inland latitudinally (Fig. 2(b)).

## Discussion

In this study, we predicted how changing climatic and land-use patterns can alter the risk for cholera infection at very fine spatial scales for the entirety of Bangladesh between the years 2015 and 2050. Using a species distribution modelling approach, we found areas with low elevation and shorter distances to surface water to be at highest risk. Areas at low elevations have greater potential for inundation from future rainfall events, which may compromise sanitation systems and increase risk for the spread of waterborne pathogens. Not only this, but projected increases in coastal vulnerability to *V. cholerae* (Escobar et al., 2015) and more frequent heavy rainfall events will also likely increase the presence of *V. cholerae* in surface waters at these elevations (Kirby et al., 2016). Low elevation areas are also likely at greater risk for infection than those of higher elevation given human settlement patterns on low-lying arable land, along rivers and other surface water. To the extent that high population density correlates with increased risk for infection, whether through increased contact with positive cases, sanitation system strain, or under-development and poverty, these areas therefore exhibit greater potential for human-to-human cholera spread (Borroto and Martinez-Piedra, 2000; Siddique et al., 1992; Root, 1997; Penrose et al., 2010).

We find that although cells of high risk (designated as having a cholera case occurrence probabilities of 0.50 and higher) cluster along the coastline predominantly in 2015, by 2050 high-risk areas expand from the coast to inland Bangladesh with all but the northwestern district of Rangpur seeing increased clusters around surface water. The overall increased risk for infection in inland Bangladesh indicates that coastal vulnerability to infection translates to increased inland infection risk. This is worrying given the predicted doubling of ENSO events in the future which will only increase coastal cholera incidence (Cai et al., 2014; Escobar et al., 2015).

While other studies have sought to develop risk maps for cholera infection and ecological presence under current climatic conditions for the entirety of Bangladesh and for future climatic conditions in strictly coastal and marine areas, our study expanded the spatial scope of predictions under a future climate scenario to include inland Bangladesh, where approximately 70% of the population lives (Ahmad, 2019). Given ongoing efforts to reduce global cholera morbidity by 90% by 2030, our study offers valuable insight into projected high-risk areas in need of continued, if not additional, public health intervention measures to reduce the burden of disease in the coming decades.

Even in the presence of infrastructural and public health advances, predictive risk mapping studies for cholera infection risk will continue to be essential in reducing the disease burden. This is because such predictions characterize a baseline set of expectations about the distribution of infection risk if future conditions resemble current circumstances. Moreover, novel cholera strains are expected to continue to arise in Bengali waters, due in part to cholera biology in the environmental reservoir. For instance, while bacteriophage niche adaptation has allowed bacteriophages to prey on *V. cholerae* infecting zooplankton in fresh and estuary water, coevolution enables *V. cholerae* to resist bacteriophage predation (Silva-Valenzuela and Camilli, 2019; Angermeyer et al., 2018). Additionally, phages can facilitate the evolution of specific toxigenic *V. cholerae* biotypes through horizontal transfer of genes associated with virulence or enhanced environmental fitness (Faruque and Mekalanos, 2012). This suggests that aquatic interactions between bacteriophages and strains of *V. cholerae* can not only select for more environmentally persistent strains, but also more virulent strains with the capacity to seed epidemics.

Climate change is likely to affect not only the distribution of waterborne diseases inland, but also socioeconomic conditions and infrastructural integrity. Thus, further modelling studies should seek to include covariates of the latter in combination with climatic variables to predict infection risk. Such models should also consider the potential for climate-associated human migration inland from vulnerable coastal regions to influence inland risk. In developing our model, we initially found the distance from each grid cell to the coast of Bangladesh to be an important variable in predicting cholera infection occurrence, with closer distances experiencing higher cholera occurrence probabilities. However, the lack of coastline projections for 2050 prevented us from including that variable in our model. Therefore, there exists a need for accurate coastline data under future climate scenarios to support robust predictive studies into disease occurrence. In addition to supporting the need for accurate sociological variable data— which is difficult to project decades into the future—remote sensing data could fill this need and in turn be useful in training models that seek to consider the interplay between human hosts and their environment on the risk for cholera infection.

As with our study, to generate valid risk predictions future models must also rely on robust case incidence data that reflects actual disease prevalence. Risk predictions from correlative models may also improve with added model complexity, but potentially at the expense of explanatory power. In future cholera infection risk forecasting studies, researchers should consider the use of hierarchical spatial models or neural networks in spatial distribution modelling that have been shown to generate robust predictions in emerging infectious disease studies (Métras et al., 2015; Redding et al., 2017; Asadgol et al., 2019; Deneu et al., 2021).

There is also a need for mechanistic models of transmission. Species distribution models (SDMs), like that of this study, represent a key first step in developing such models, but may not include the effect of climate-sensitive ecological processes on model predictions (Cuddington et al., 2013). Therefore, in the context of global change, modelling the spatial distribution of risk for cholera infection is best done using process-based models that will use our model’s infection probabilities, consider the correlative components of our model, and incorporate the ecological mechanisms influencing the distribution of cholera and human transmission. Nevertheless, our study holds importance in providing robust inland climate-associated cholera infection risk predictions that can inform preventive Bengali public health strategies.

## Data Availability

All code used in the analysis is publicly available on github (github.com/sophiakruger/cholera_risk) and released under the GNU Public License v.3 (Stallman, 2007). This paper uses publicly available data sets (obtained from https://github.com/HopkinsIDD/Bangladesh-Cholera-Serosurvey/tree/master/data). These data are de-identified and intended for public use.

github.com/sophiakruger/cholera_risk

## Acknowledgements

This research was made possible in part by a Sustainability Scholars grant awarded by the Undergraduate Research Opportunities Program at the University of St. Thomas. The authors acknowledge the Minnesota Supercomputing Institute (MSI) at the University of Minnesota for providing resources that contributed to the research results reported within this paper. URL: http://www.msi.umn.edu. The authors also thank Charlie Frye for his helpful recommendations regarding covariate data.

## Supplementary Materials

### S1. Spatial Data Manipulation in R and ArcGIS

To interface R programming language with ArcGIS Pro, a Geographic Information Systems (GIS) software, we used the ‘arcgisbinding’ package in R to facilitate loading ArcGIS raster layers into R and exporting raster layers from R to ArcGIS (ESRI, 2019). All raster data available from source as TIFF files were uploaded into ArcGIS Pro, projected to WGS84, resampled to a resolution of 250m square grid cells, and cropped to the rectangular extent surrounding the country of Bangladesh. We used the administrative boundary level 0 provided by the GADM spatial database (v. 3.6) as the extent for our study area (88.01057°W, 92.67366°E, 20.74111°S, 26.63407°N). Image service layers, provided by ESRI’s Living Atlas Portal, were imported into ArcGIS through the portal, then projected, resampled, and cropped with the same procedure as described for all TIFF raster files.

Of the covariates used for 2015, the average precipitation and temperature raster layers were products of additional data manipulation that occurred in R and ArcGIS Pro. For 2050, the precipitation, temperature, and elevation rasters required additional manipulation. Below we detail additional steps taken to include these data as spatial covariates in our model.

#### S1.1 Average Precipitation and Temperature Rasters (2015 and 2050)

The average precipitation and maximum and minimum temperature rasters were created from average monthly climate data (as NetCDF files) provided by TerraClimate (Abatzoglou, et al., 2018). The NetCDF files were converted in R to GeoTIFF files (see “file-coversion.R” in repository), then exported to ArcGIS for projecting, resampling, and cropping using the ModelBuilder functionality. Using the raster calculator tool in ModelBuilder, the raster bands for October 2015 through January 2016 were averaged to estimate the average monthly precipitation accumulation (mm), maximum temperature (°C), and minimum temperature (°C) experienced in Bangladesh during the survey period of the Azman data. The months were selected with the assumption that the climatic conditions during the survey period would have influenced measured incidence. To consider the possibility that cholera incidence is a lagging indicator predicted by rainfall and temperatures of the monsoon season, we also created separate monsoon temperature and precipitation covariates for 2015 using the same methodology to average the monthly values for June through September. This process of averaging the precipitation and temperature data in ArcGIS was also replicated in R (see “2015-raster-manipulation.R” in repository) to ensure that estimated averages were consistent across platforms. For 2050, the average precipitation and maximum and minimum temperature rasters from WorldClim (v2.1) were created following the same procedure as described for the TerraClimate files, though these layers were downloaded from source as TIFF files and thus did not need conversion (see “2050-raster-manipulation.R” for the averaging process in R).

#### S1.2 Elevation Raster (2050)

The 90-meter coastal elevation layer from Kulp and Strauss (2018) at ClimateCentral was projected, resampled, and cropped to Bangladesh in ArcGIS. The raster layer was then exported to R wherein missing values were imputed from the 2015 elevation layer using a generalized linear model in R (see “2050-raster-manipulation.R” in repository).

Once all layers were manipulated using the ModelBuilder functionality in ArcGIS, all raster layers were exported for use in R. To avoid memory overload issues in using R solely to project, resample, and crop global raster layers, our methodology relies on ArcGIS’s ModelBuilder to do the same and export all raster layers to R for analysis. Nonetheless, the code written in R follows the procedure streamlined in ArcGIS (e.g., projecting, resampling, and cropping) to encourage reproducibility and to also ensure that all raster layers, once imported into R, align to the desired raster cell size, global projection, and study area extent (see “2015-raster-manipulation.R” and “2050-raster-manipulation.R” in repository).

**Table S1.**
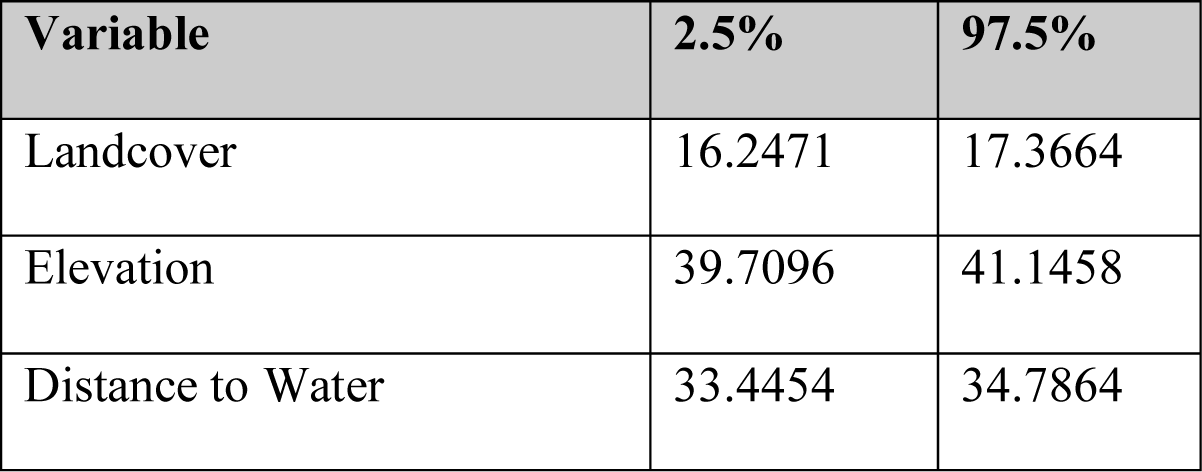
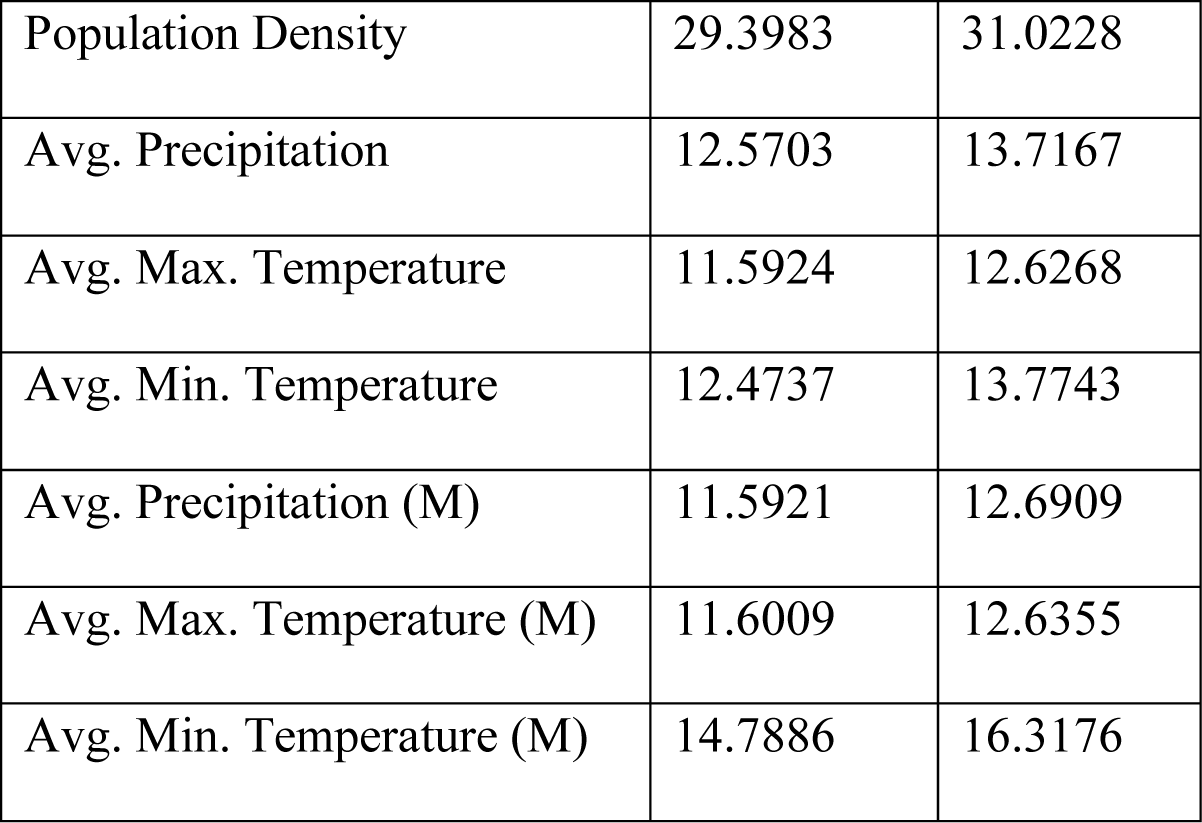
Confidence interval (95%) of the variable importance (mean decrease in Gini) for each covariate included in our full model. A higher mean decrease in Gini coefficient reflects a variable’s greater importance to the random forest model. The (M) differentiates our temperature and precipitation variables between data taken during the monsoon period (M) and not.

**Table S2.**
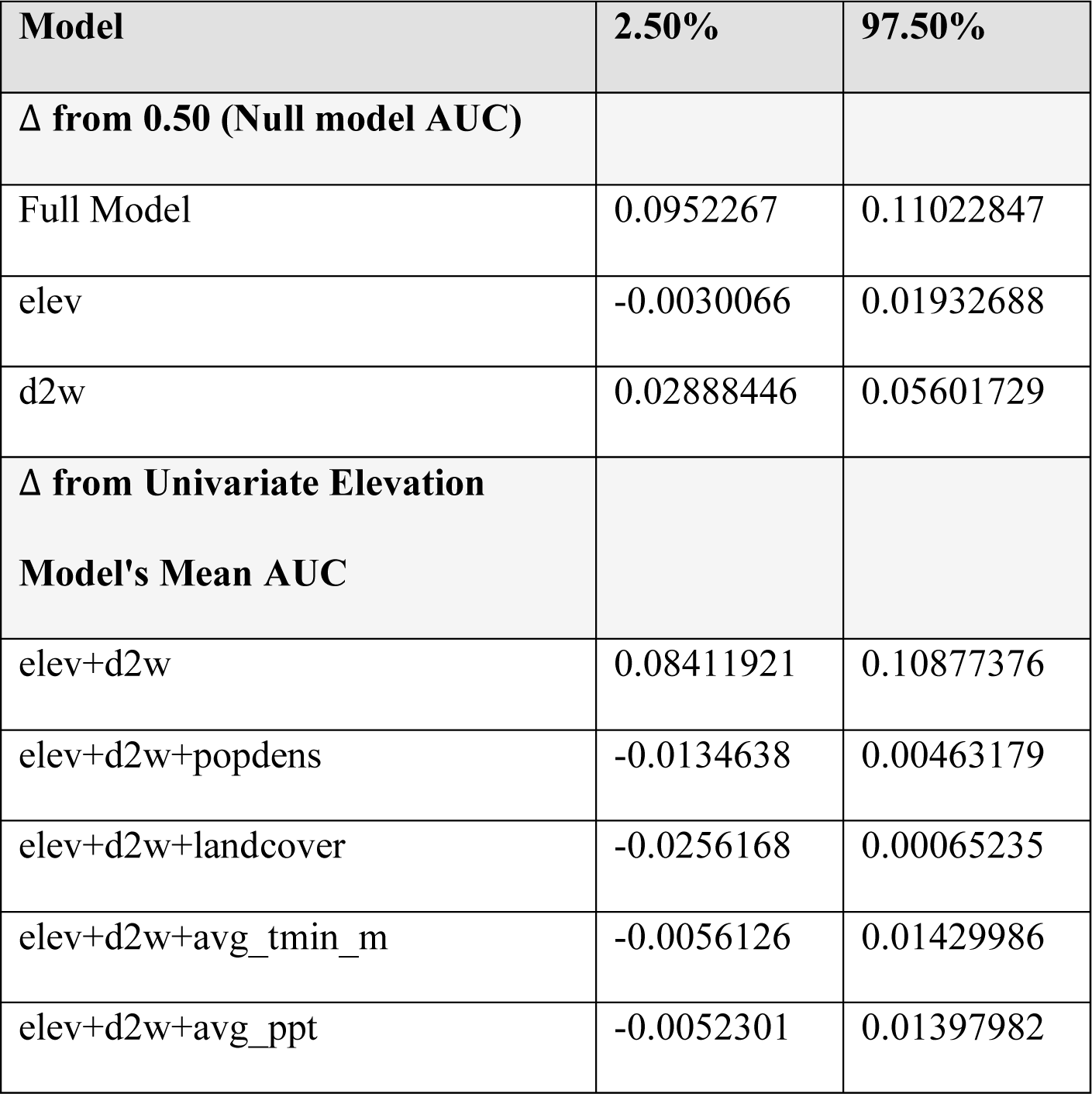
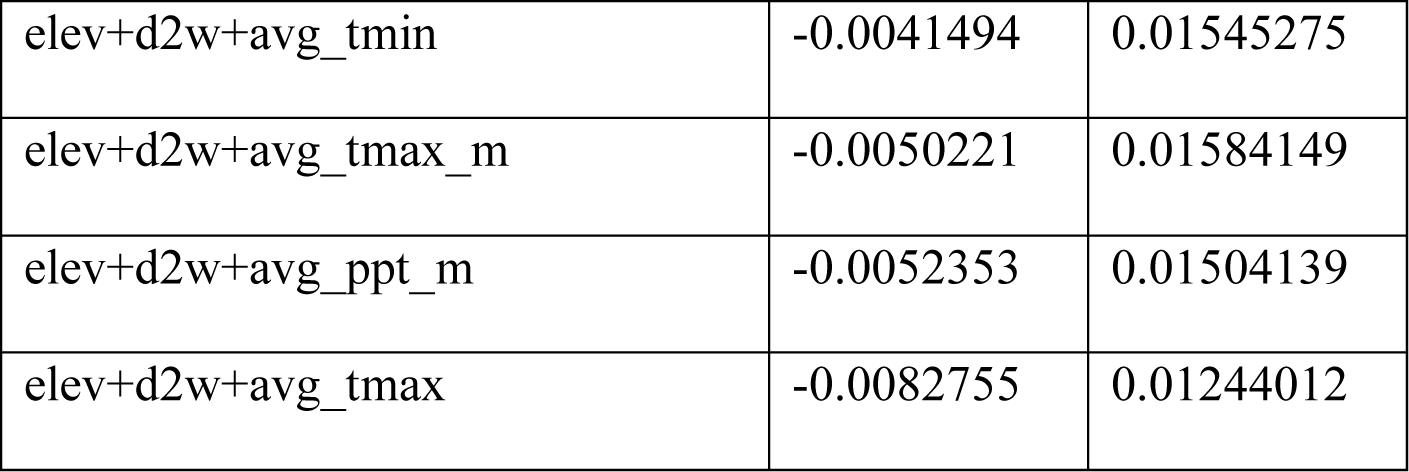
Change in AUC confidence interval for each model obtained in our stepwise model selection process.

**Figure S1.**
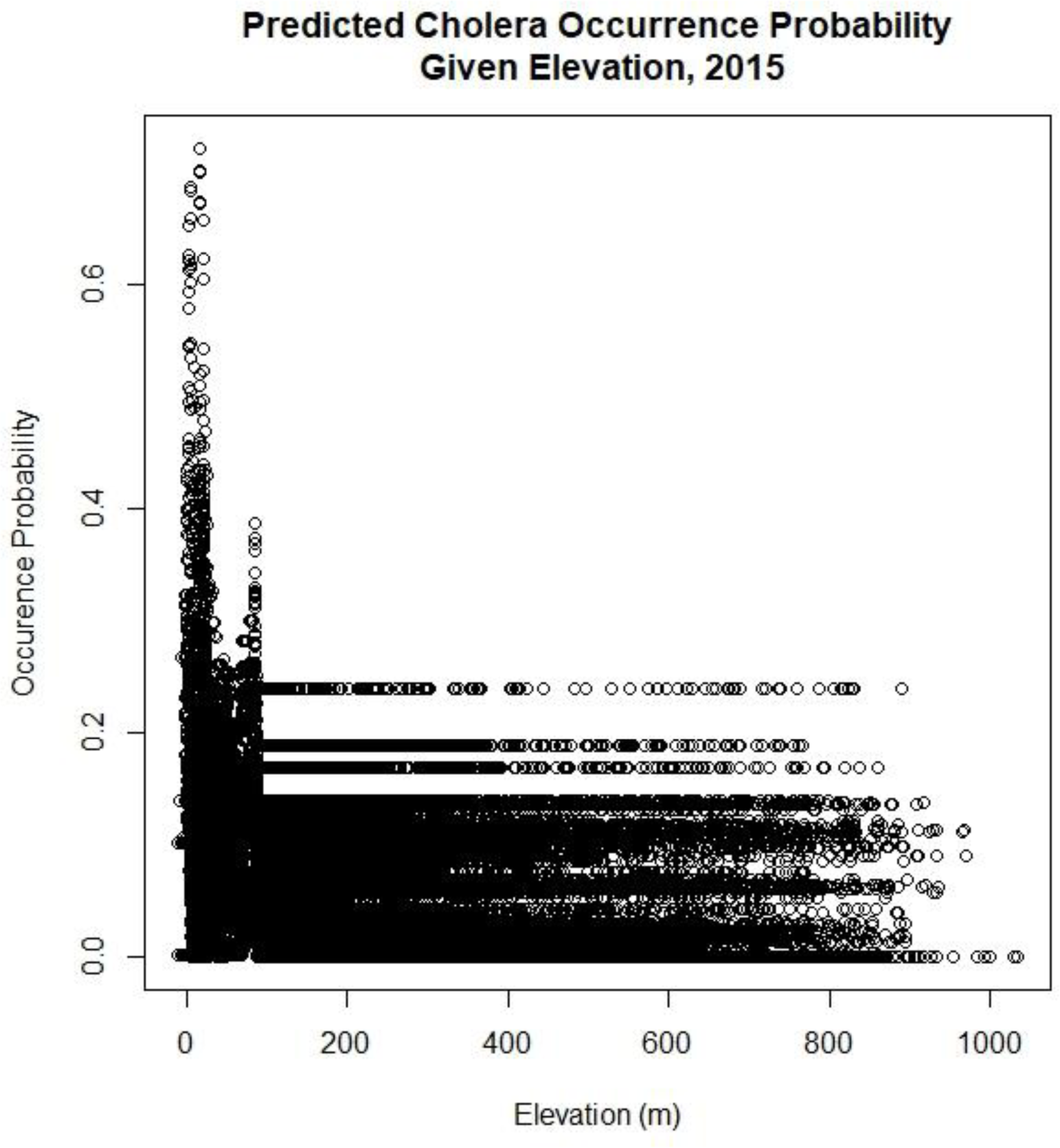
Response curve of the 2015 occurrence probabilities generated by our bivariate random forest model against the elevation values for 2015.

**Figure S2.**
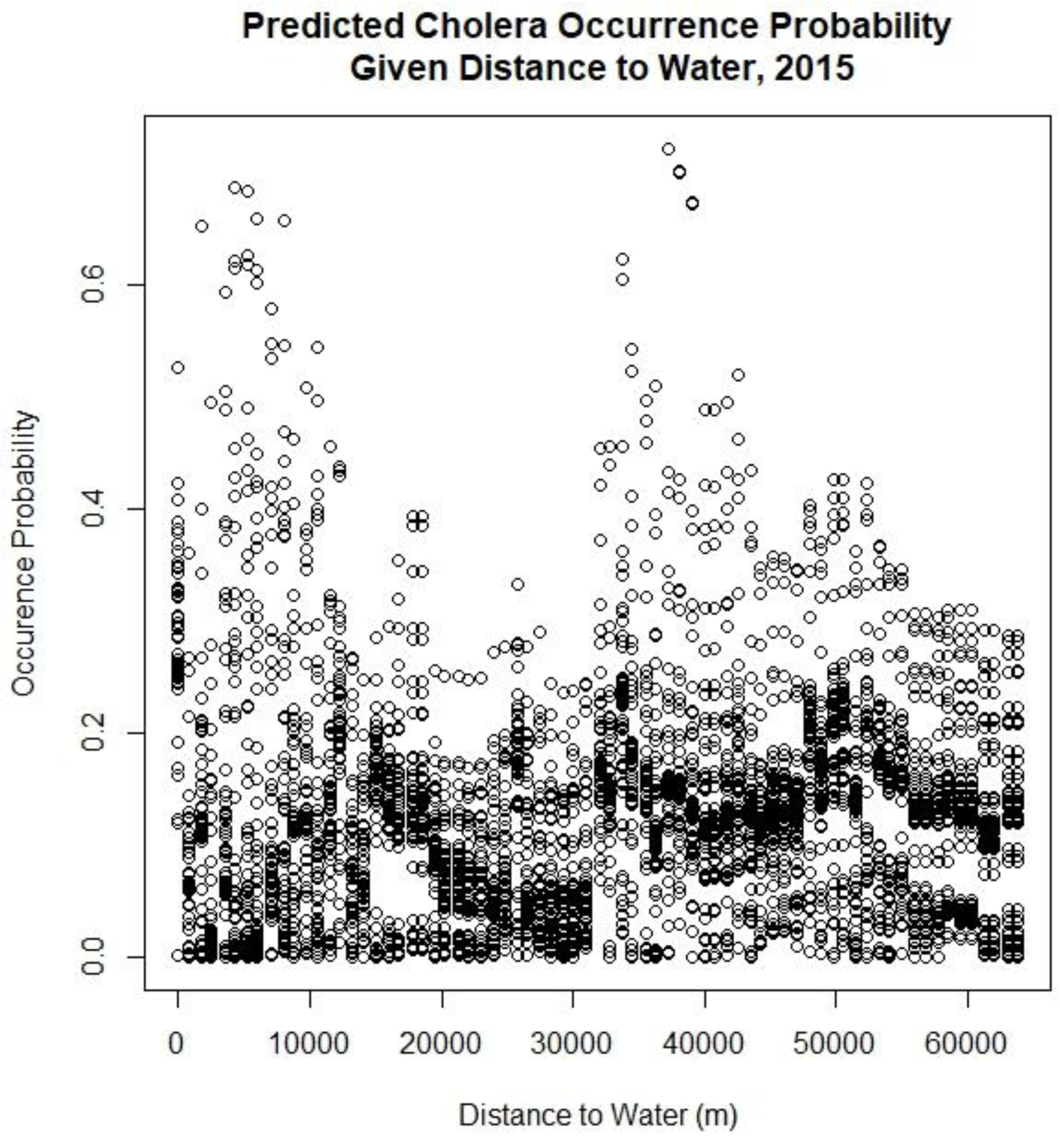
Response curve of the 2015 occurrence probabilities generated by our bivariate random forest model against the distance to water values for 2015.

**Figure S3.**
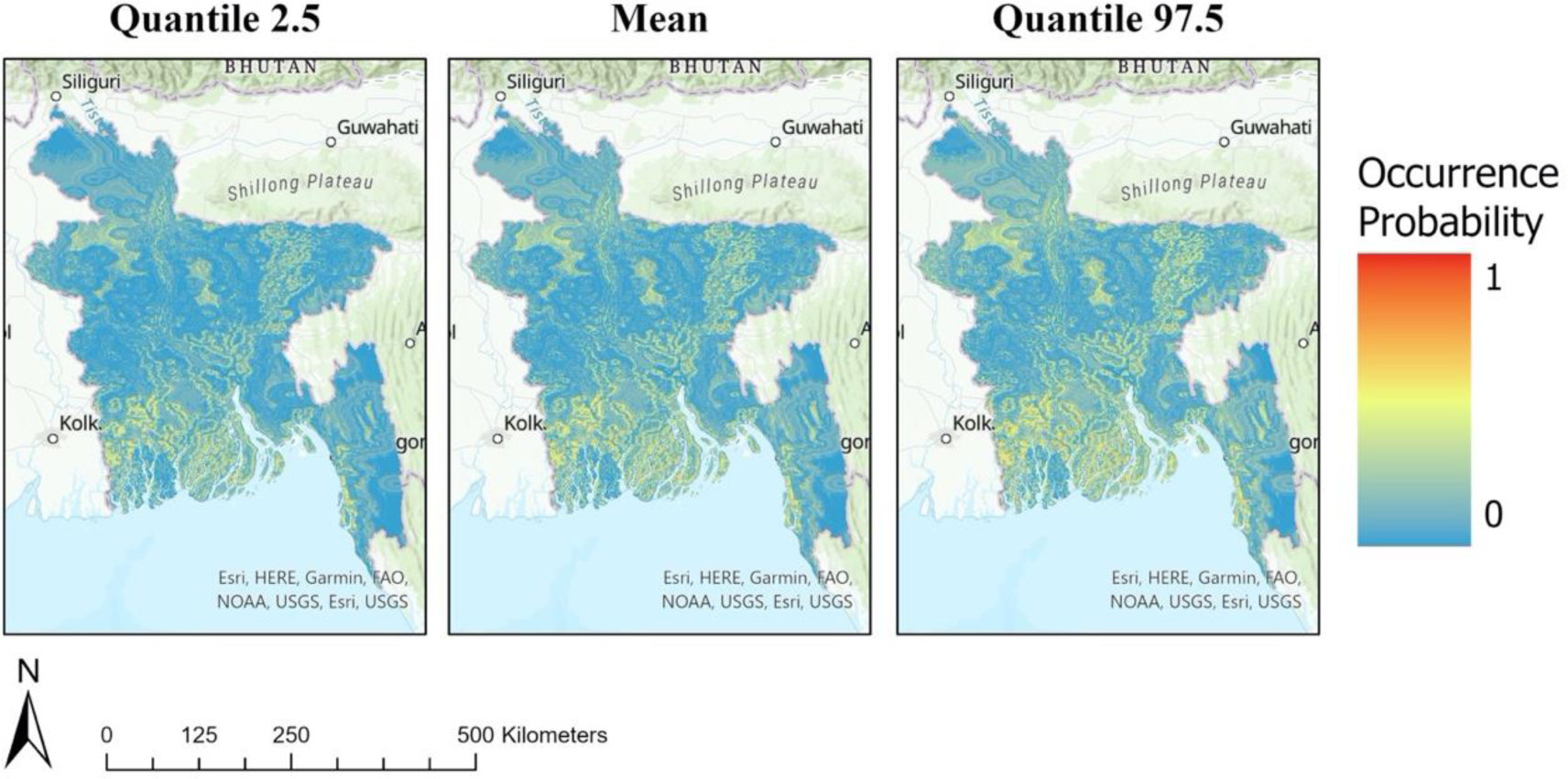
Mean and quantile (2.5% and 97.5%) risk maps predicting cholera infection risk for 2015 from predictions constructed by our best-fitting random forest model. Risk values range from 0 to 1 with 1 representing the highest risk for cholera infection in the specified geographic area.

**Figure S4.**
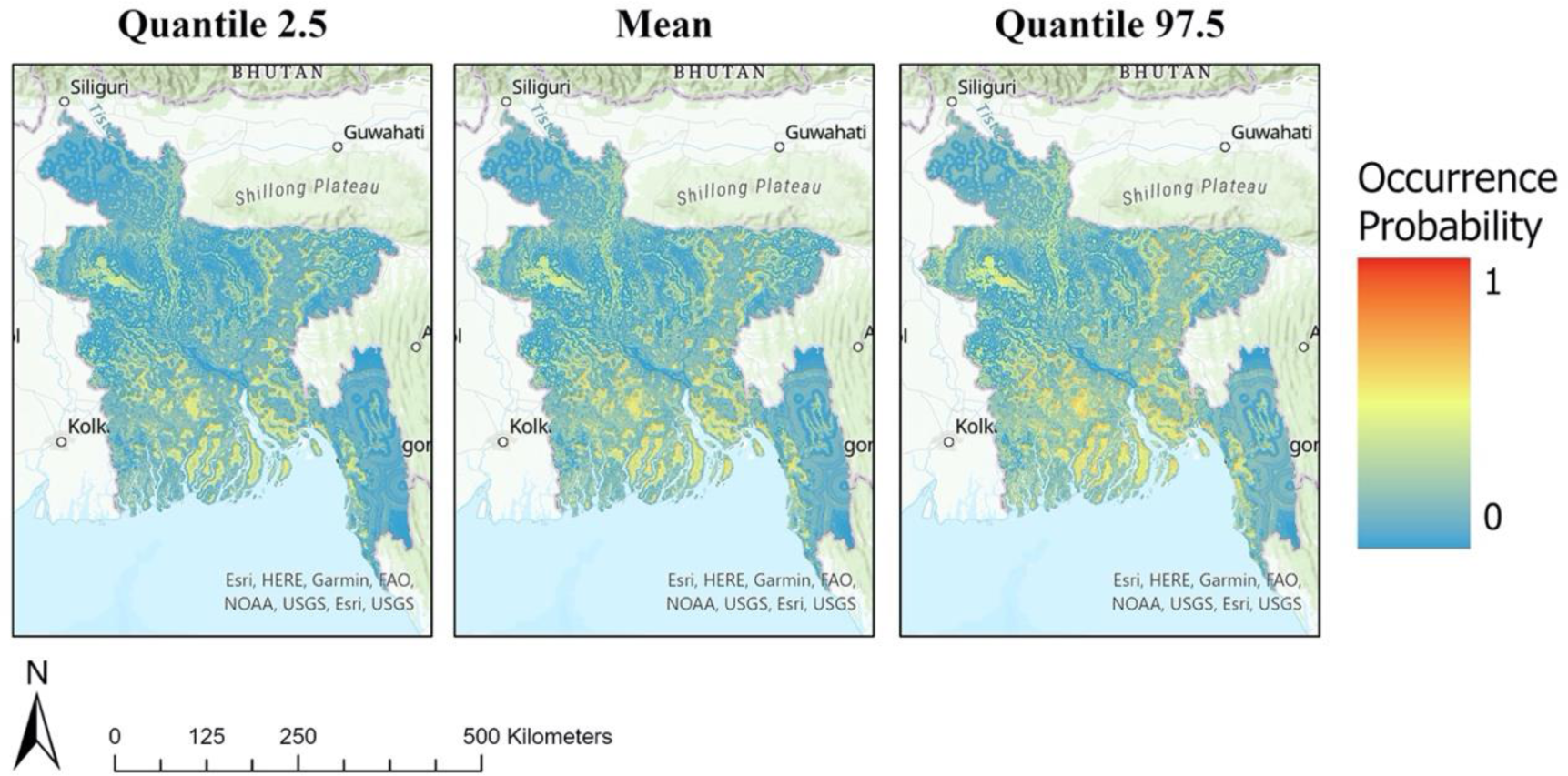
Mean and quantile (2.5% and 97.5%) risk maps predicting cholera infection risk into 2050 from predictions constructed by our best-fitting random forest model. Risk values range from 0 to 1 with 1 representing the highest risk for cholera infection in the specified geographic area.

